# Neoadjuvant Immunotherapy and Chemoradiation Followed by Esophagectomy for Esophageal Cancer

**DOI:** 10.1101/2023.10.01.23296396

**Authors:** Nikhil Panda, Beverly Fu, Alexandra Potter, Hugh G. Auchincloss, Samuel J. Klempner, Michael Lanuti, Chi-Fu Yang, Lana Schumacher

## Abstract

**Background:** Treatment of locally advanced esophageal cancer includes neoadjuvant chemoradiation (chemoRT) and esophagectomy. We evaluated perioperative and oncologic outcomes among patients who received neoadjuvant chemoRT and immunotherapy (I/O).

**Methods:** Adults who underwent esophagectomy following neoadjuvant chemoRT or chemoRT+I/O for T1-4, N0-3, M0 esophageal cancer were identified from the National Cancer Database (2012-2020). Unadjusted, propensity score-matched, and Cox proportional hazards analyses compared perioperative outcomes and three-year overall survival (OS) between neoadjuvant chemoRT versus chemoRT+I/O cohorts.

**Results:** Among 17,937 patients, 261 (1.5%) received neoadjuvant chemoRT+I/O. ChemoRT+I/O patients were younger (62 versus 64 years, p=0.002) and had a longer interval between chemotherapy and surgery (104.5 versus 97.0 days, p<0.001) compared with chemoRT patients. Among the chemoRT+I/O cohort, there were more undifferentiated tumors (46.4% versus 34.3%, p<0.001) with adenocarcinoma histology (93.9% versus 81.2%, p<0.001) compared with the chemoRT cohort. On unadjusted analysis, there were no significant differences regarding margin positivity, 30-day readmission, 30-day mortality, or 90-day mortality. ChemoRT+I/O patients had higher 3-year OS (61.4% 95%CI [54.2-67.7] versus 55.1% [54.3-55.9], p=0.02), more lymph nodes resected (median 17.0 IQR [11.0-25.0] versus 15.0 [10.0-22.0], p=0.007), and less pathologic nodal downstaging from N2 to N1/N0 (36.8% vs 48.4%, p<0.001) than chemoRT patients. Propensity score-matched analyses (n=217) revealed no differences in perioperative outcomes and 3-year OS (65.0% 95%CI [57.1-71.8] versus 56.0% [48.0-63.3], p=0.11) between the chemoRT+I/O and chemoRT cohort.

**Conclusions:** There were similar perioperative outcomes and 3-year OS between patients who received neoadjuvant chemoRT+I/O and chemoRT, supporting the feasibility of adding immunotherapy to neoadjuvant regimens.

## INTRODUCTION

The landmark Chemoradiotherapy for Oesophageal Cancer Followed by Surgery Study (CROSS) established trimodality therapy as a standard of care for patients with locally advanced, resectable esophageal cancer^1^. Despite this multimodal treatment approach, patients remain at high risk of local or distant recurrence^2^. As such, approaches to improve upon micrometastatic control are needed.

Immune checkpoint inhibitors (I/O) are an attractive strategy to potentially improve upon chemoradiotherapy (chemoRT). I/O, in the presence of an intact primary tumor, may enhance expansion of tumor-specific CD8+ T-cell populations.^3^ Additionally, the tumor and microenvironment remodeling that occurs with chemoRT may enhance the efficacy of chemoRT, translating into improved pathologic outcomes. Small pilot trials and retrospective analyses point toward preserved surgical morbidity with the addition of I/O, especially in esophageal squamous cell carcinoma^4^. For example, the FRONTiER JCOG1804E^5^ trial demonstrated that neoadjuvant chemotherapy and I/O followed by surgery was well-tolerated with efficacy potential in esophageal cancer squamous cell carcinoma (ESCC). The additional short- and intermediate-term efficacy of adding I/O to neoadjuvant chemoRT regimens, especially for locally advanced esophageal adenocarcinoma, represents an ongoing knowledge gap.

The purpose of this study was to evaluate the perioperative and oncologic outcomes of patients who underwent neoadjuvant chemoRT+I/O and esophagectomy for locally advanced esophageal cancer in a large national database. Based on preliminary data from phase I and II clinical trials,^6^, as well as the use of neoadjuvant immunotherapy for other solid tumors, we hypothesized that outcomes would be comparable among patients who received neoadjuvant chemoRT+I/O compared with chemoRT alone. If confirmed, these results would provide foundational data to support the feasibility and safety of novel I/O-based neoadjuvant approaches.

## PATIENTS AND METHODS

### Data Source

The protocol for this study was approved by the Massachusetts General Brigham Institutional Review Board and followed the strengthening of reporting of observational studies in epidemiology (STROBE Checklist, Supplement 1). Patients in the National Cancer Database (NCDB) diagnosed from 2012-2020 with clinical T1-4, N0-3, M0 esophageal cancer who underwent esophagectomy after chemoRT or chemoRT+I/O were identified using International Classification of Diseases for Oncology (3^rd^ edition, ICD-O-3) histology and topography codes (Supplement 2). The 2012-2020 study period was selected as neoadjuvant I/O was rarely used before 2012 and the NCDB is updated through 2020. Patients were excluded if diagnosed with M1 disease or preoperative treatment consisted of radiation without chemotherapy.

### Exposure and Outcome Measures

Patients were organized into cohorts according to the neoadjuvant therapy received: chemoRT or chemoRT+I/O. Patients were included if neoadjuvant chemoRT commenced six months and I/O seven months prior to surgery. The primary endpoint was 3-year overall survival (OS) measured from time of diagnosis to death or last follow-up.

### Statistical Analysis

Continuous data were summarized using means and standard deviations or median and interquartile ranges. Categorical data were summarized using frequencies and percentages. Differences in baseline characteristics and unadjusted outcomes between chemoRT and chemoRT+I/O cohorts were assessed using Student’s *t*-test or Wilcoxon rank-sum tests (continuous) and the Pearson’s chi-square or Fisher’s exact tests (categorical variables). Factors associated with receipt of neoadjuvant chemoRT+I/O were identified using the lasso penalized logistic regression estimator, a form of supervised machine learning. The *a priori* model variables were age, sex, race, income, education, year of diagnosis, and distance from hospital. Overall survival rates were analyzed using the log-rank test. A multivariable Cox proportional hazards model compared survival between the chemoRT and chemoRT+I/O cohorts, adjusting for age, sex, race, Charlson comorbidity (CDCC) score, clinical T and N stages, insurance status, facility type, income, education, grade, tumor location, histology, year of diagnosis, distance from hospital, and operative approach (ie open, video-assisted, or robot-assisted esophagectomy) was performed. The proportional hazards assumption was tested for the Cox model using smooth scaled Schoenfeld residual plots; no violations were found. Linearity was confirmed for all continuous predictors encompassed in the Cox regression analysis using Martingale residuals.

Differences in outcomes between the chemoRT versus chemoRT+I/O cohorts were further assessed using a propensity score-matched analysis. Propensity scores were developed, defined as the probability of treatment with chemoRT versus chemoRT+I/O, conditional on *a priori* clinically relevant variables: age, sex, race, CDCC score, median census-tract education and income levels, clinical T and N stages, tumor location, insurance type, histology, grade, facility type, open versus minimally invasive approach, and year of diagnosis. The most appropriately matched pairs were chosen using a greedy nearest neighbor algorithm without replacement with a caliper of 0.01 on the propensity score^7^. Balance of the match was evaluated using standardized differences. Statistical significance was defined as a two-sided alpha≤0.05. Analyses were performed using STATA (version 16, StataCorp, College Station, TX, USA).

## RESULTS

Among 17,937 patients who underwent esophagectomy for T1-4, N0-3, M0 esophageal cancer between 2012 and 2020, 17,676 (98.5%) received neoadjuvant chemoRT and 261 (1.5%) received neoadjuvant chemoRT+I/O. The inclusion of neoadjuvant I/O in neoadjuvant regimens increased from 2012 (0.1%) to 2020 (2.2%).

### Patient and Disease Characteristics

Baseline demographic information, clinicopathologic characteristics, and procedure-specific details are shown in Tables 1-2. Compared with patients who received neoadjuvant chemoRT, chemoRT+I/O patients were younger (median 62.0 [IQR 55.0-69.0] versus 64.0 [57.0-70.0] years, p=0.002), male (88.1% versus 83.1%, p=0.03), with undifferentiated tumors (46.4% versus 34.3%, p<0.001), adenocarcinoma histology (93.9% versus 81.2%, p<0.001), and received treatment at an academic research program (55.5% versus 46.2%, p=0.003). ChemoRT+I/O patients had a longer interval between chemotherapy and surgery (median 104.5 [IQR 87.0-127.0] versus 97.0 [IQR 84.0-114.0] days, p<0.001). There were no differences in interval between neoadjuvant radiation and surgery between cohorts, nor operative approach (e.g., open, video-assisted, or robot-assisted esophagectomy). Results from the lasso analysis demonstrated that younger age (OR 0.97 95%CI [0.94-0.99], p=0.02) and longer intervals between chemotherapy and surgery (OR 1.02 95%CI[1.01-1.02],p<0.01) were associated with receipt of I/O (Supplementary Table 1). In this cohort, patients with squamous cell histology had lower odds of receiving chemoRT+I/O (OR 0.43 95%CI[0.20-0.91], p=0.03).

**Table 1:**
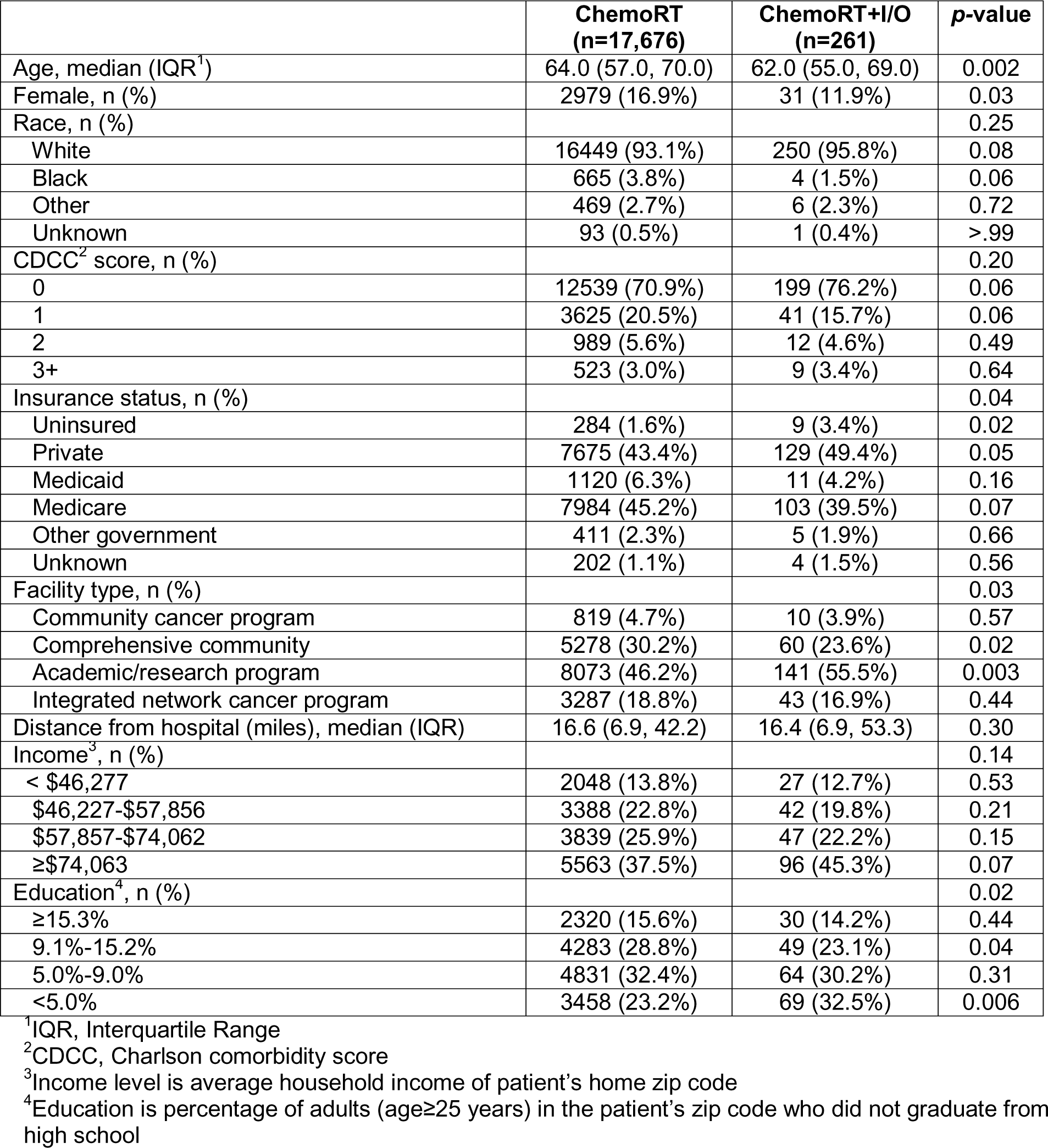
Baseline Characteristics of Patients who Received Neoadjuvant ChemoRT or ChemoRT+I/O.

**Table 2:** Disease Characteristics of Patients who Received Neoadjuvant ChemoRT or ChemoRT+I/O.

|  | **ChemoRT**<br>**(n=17,676)** | **ChemoRT+I/O**<br>**(n=261)** | **p-value** |
| --- | --- | --- | --- |
| Clinical T Status, n (%) |  |  | 0.52 |
| T0 | 6 (<1%) | 0 (0.0%) | >.99 |
| T1 | 721 (4.1%) | 7 (2.7%) | 0.26 |
| T2 | 3424 (19.4%) | 55 (21.1%) | 0.49 |
| T3 | 11905 (67.4%) | 176 (67.4%) | 0.98 |
| T4 | 339 (1.9%) | 7 (2.7%) | 0.37 |
| Clinical N Status, n (%) |  |  | 0.05 |
| N0 | 6677 (37.8%) | 80 (30.7%) | 0.02 |
| N1 | 7834 (44.3%) | 130 (49.8%) | 0.07 |
| N2 | 2497 (14.1%) | 46 (17.6%) | 0.11 |
| N3 | 372 (2.1%) | 4 (1.5%) | 0.52 |
| Tumor size (mm), median (IQR)<sup>2</sup> | 40.0 (28.0, 60.0) | 46.0 (30.0, 60.0) | 0.16 |
| Tumor location, n (%) |  |  | 0.005 |
| Upper third of esophagus | 199 (1.1%) | 1 (0.4%) | 0.26 |
| Middle third of esophagus | 1867 (10.6%) | 14 (5.4%) | 0.007 |
| Lower third of esophagus | 14165 (80.1%) | 228 (87.4%) | 0.004 |
| Overlapping lesion | 763 (4.3%) | 5 (1.9%) | 0.06 |
| Unspecified location | 682 (3.9%) | 13 (5.0%) | 0.35 |
| Histology, n (%) |  |  | <.001 |
| Adenocarcinoma | 14348 (81.2%) | 245 (93.9%) | <.001 |
| Squamous | 2802 (15.9%) | 12 (4.6%) | <.001 |
| Other | 526 (3.0%) | 4 (1.6%) | <.001 |
| Grade, n (%) |  |  | <.001 |
| Well differentiated | 472 (2.7%) | 4 (1.5%) | 0.25 |
| Moderately differentiated | 4307 (24.4%) | 74 (28.4%) | 0.14 |
| Poorly differentiated | 5095 (28.8%) | 42 (16.1%) | <.001 |
| Undifferentiated, anaplastic | 6069 (34.3%) | 121 (46.4%) | <.001 |
| Unknown | 1683 (9.5%) | 20 (7.7%) | 0.30 |
| Interval: chemotherapy to surgery (days), median (IQR) | 97.0 (84.0, 114.0) | 104.5 (87.0, 127.0) | <.001 |
| Interval: radiation to surgery (days), median (IQR) | 95.0 (84.0, 111.0) | 95.5 (81.0, 111.0) | 0.89 |
| Interval: I/O and surgery (days), median (IQR) | - | 102.0 (86.0, 125.0) | - |
| Operative Approach, n (%) |  |  | 0.08 |
| Open | 6702 (37.9%) | 98 (37.5%) | 0.90 |
| VATS | 4258 (24.1%) | 80 (30.7%) | 0.01 |
| Robot | 2243 (12.7%) | 27 (10.3%) | 0.26 |

### Clinical and Histopathologic Outcomes

Table 3 shows unadjusted clinicopathologic outcomes. The chemoRT and chemoRT+I/O cohorts did not differ in terms of pathologic stage, complete pathologic response, margin positivity, 30-day readmission, 30- or 90-day mortality. The chemoRT+I/O cohort had more lymph nodes examined (median 17.0 [IQR 11.0-25.0] versus 15.0 [10.0-22.0], p=0.007) compared with chemoRT and a lower rate of pathologic nodal downstaging (N2 to N1 or N0: 36.8% versus 48.4%, p<0.001) On unadjusted analysis, the chemoRT+I/O patients appeared to experience higher 3-year OS (61.4% 95%CI[54.2-67.7] versus 55.1% 95%CI[54.3-55.9], p=0.02) than the chemoRT cohort (Figure 1A). Additional non-propensity-matched patient and disease-specific factors associated with overall survival are shown in Supplementary Table 2.

**Table 3:** Unadjusted Clinicopathologic Outcomes for Patients who Received ChemoRT or ChemoRT+I/O.

|  | **ChemoRT**<br>**(n=17,676)** | **ChemoRT+I/O**<br>**(n=261)** | **p-value** |
| --- | --- | --- | --- |
| Pathologic T Stage, n (%) |  |  | 0.64 |
| T0 | 2777 (15.7%) | 35 (13.4%) | 0.31 |
| T1 | 1950 (11.0%) | 17 (6.5%) | 0.02 |
| T2 | 1910 (10.8%) | 20 (7.7%) | 0.10 |
| T3 | 3765 (21.3%) | 46 (17.6%) | 0.15 |
| T4 | 70 (0.4%) | 1 (0.4%) | >.99 |
| Pathologic N Stage, n (%) |  |  | 0.77 |
| N0 | 6947 (39.3%) | 75 (28.7%) | 0.001 |
| N1 | 2381 (13.5%) | 31 (11.9%) | 0.45 |
| N2 | 1097 (6.2%) | 14 (5.4%) | 0.58 |
| N3 | 409 (2.3%) | 4 (1.5%) | 0.53 |
| Pathologic M Stage, n (%) |  |  | >.99 |
| M0 | 3 (<1%) | 0 (0.0%) | >.99 |
| M1 | 87 (0.5%) | 1 (0.4%) | >.99 |
| Complete pathologic response, n (%) | 2308 (22.4%) | 29 (24.8%) | 0.54 |
| Nodal Upstage<sup>1</sup>, n (%) | 1772 (26.5%) | 23 (28.8%) | 0.49 |
| Nodal Downstage: N2 to N0<sup>2</sup>, n (%) | 848 (34.0%) | 11 (23.9%) | 0.18 |
| Nodal Downstage: N2 to N1 or N0<sup>2</sup>, n (%) | 1208 (48.4%) | 17 (36.8%) | <0.001 |
| Patients with Lymph Nodes Examined, n (%) | 16691 (94.4%) | 247 (94.6%) | 0.88 |
| Lymph Nodes Examined, median (IQR) | 15.0 (10.0, 22.0) | 17.0 (11.0, 25.0) | 0.007 |
| Surgical margins, n (%) |  |  | 0.94 |
| No residual tumor | 16288 (92.1%) | 241 (92.3%) | 0.91 |
| Residual tumor, not specified | 463 (2.6%) | 7 (2.7%) | 0.95 |
| Microscopic residual tumor | 457 (2.6%) | 6 (2.3%) | 0.77 |
| Macroscopic residual tumor | 31 (0.2%) | 1 (0.4%) | 0.43 |
| Unknown or not evaluable | 437 (2.5%) | 6 (2.3%) | 0.42 |
| Margin positivity, n (%) | 951 (5.4%) | 14 (5.4%) | 0.99 |
| Length of stay (days), median (IQR) | 9.0 (7.0, 13.0) | 9.0 (7.0, 13.0) | 0.90 |
| 30-day unplanned readmission, n (%) | 942 (5.3%) | 13 (5.0%) | 0.82 |
| 30-day mortality<sup>3</sup>, n (%) | 445 (2.5%) | 4 (1.5%) | 0.25 |
| 90-day mortality<sup>3</sup>, n (%) | 1070 (6.1%) | 11 (4.2%) | 0.63 |

<sup>1</sup>Denominator is the number with cN0 disease (chemoRT: n=6677; chemoRT+I/O: n=80)
<sup>2</sup>Denominator is the number with cN2 disease (chemoRT: n=2497; chemoRT+I/O: n = 46)
<sup>3</sup>Mortality data is unavailable for 2018 and contributes to the majority of unknown 30- and 90-day mortality values; percentages are calculated out of known data

**Figure 1:**
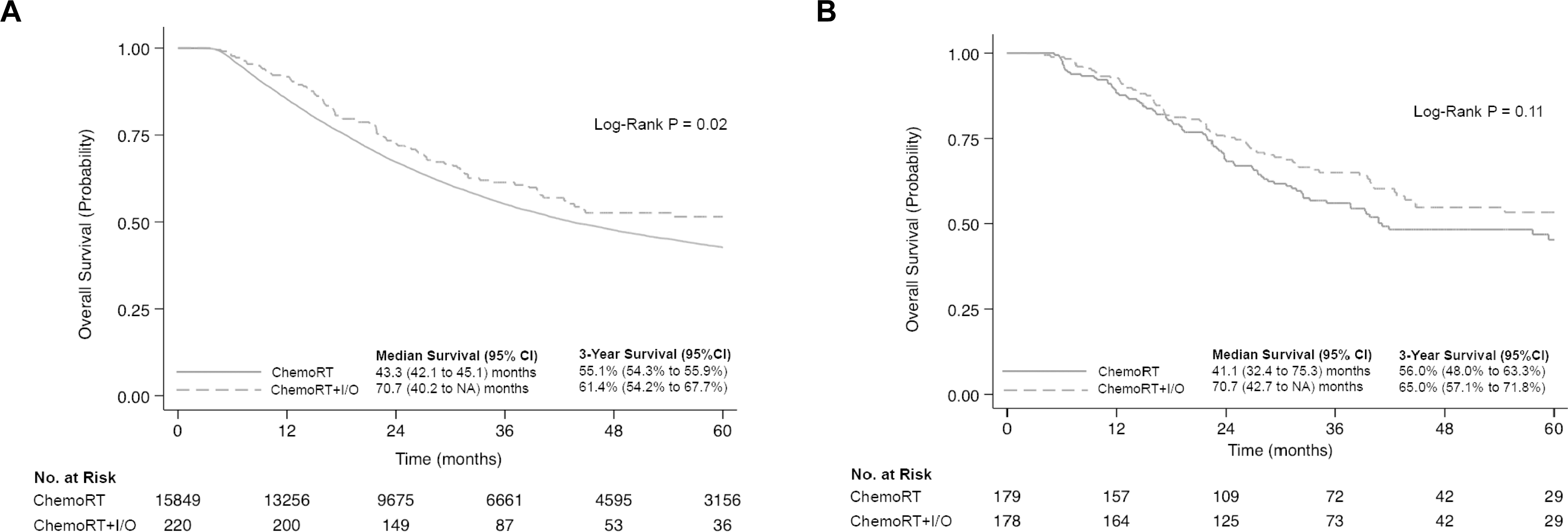
Survival for Patients who Received Neoadjuvant ChemoRT or ChemoRT+I/O. (A) Overall survival analysis, excluding n=1827 and n=41 missing patients in chemoRT and chemoRT+I/O cohorts, respectively. (B) Propensity-matched overall survival analysis, excluding n=38 and n=39 missing patients in chemoRT and chemoRT+I/O cohorts, respectively.

Propensity-matched cohorts (n=217) were well matched by demographics, clinicopathologic characteristics, and procedure-specific details (Tables 4-5). There were no significant differences in clinicopathologic outcomes, including nodes examined (median 17.0 [IQR 11.0-26.0] versus 17.0 [12.0-24.0], p = 0.98) or rate of pathological nodal downstaging (N2 to N1 or N0: 32.5% versus 39.5%, p=0.45). There were also no significant differences in 30-day mortality (0.9% versus 0.5%, p=0.88) and 90-day mortality (3.7% versus 5.1%, p=0.73) when comparing propensity-matched chemoRT+I/O versus chemoRT cohorts (Table 6). Three-year OS was also similar between patients receiving neoadjuvant chemoRT+I/O versus chemoRT (65.0% 95%CI [57.1-71.8] versus 56.0% 95%CI [48.0-63.3]), p=0.11, Figure 1B).

**Table 4:** Baseline Characteristics for Patients who Received Neoadjuvant ChemoRT or ChemoRT+I/O – Propensity Matched Analysis.

|  | **ChemoRT**<br>**n=217** | **ChemoRT+I/O**<br>**n=217** | **Standardized**<br>**Differences** |
| --- | --- | --- | --- |
| Age, median (IQR)<sup>1</sup> | 63.0 (56.0, 68.0) | 62.0 (56.0, 69.0) | -4.8 |
| Female, n (%) | 24 (11.1%) | 24 (11.1%) | 0.0 |
| Race, n (%) |  |  |  |
| White | 214 (98.6%) | 210 (96.8%) | -8.5 |
| Black | 1 (0.5%) | 3 (1.4%) | 5.7 |
| Other | 2 (0.9%) | 4 (1.8%) | 6.1 |
| CDCC<sup>2</sup> score, n (%) |  |  |  |
| 0 | 174 (80.2%) | 165 (76.0%) | -1.1 |
| 1 | 29 (13.4%) | 32 (14.7%) | -3.6 |
| 2 | 7 (3.2%) | 11 (5.1%) | 8.3 |
| 3+ | 7 (3.2%) | 9 (4.1%) | 5.2 |
| Insurance status, n (%) |  |  |  |
| Uninsured | 5 (2.3%) | 5 (2.3%) | 0.0 |
| Private | 111 (51.2%) | 111 (51.2%) | 0.0 |
| Medicaid | 4 (1.8%) | 8 (3.7%) | 9.2 |
| Medicare | 95 (43.8%) | 89 (41.0%) | -5.6 |
| Other Government | 2 (0.9%) | 4 (1.8%) | 6.3 |
| Facility type, n (%) |  |  |  |
| Community cancer program | 6 (2.8%) | 6 (2.8%) | 0.0 |
| Comprehensive community | 61 (28.1%) | 55 (25.3%) | -6.3 |
| Academic/research program | 115 (53.0%) | 119 (54.8%) | 3.7 |
| Integrated network cancer program | 35 (16.1%) | 37 (17.1%) | 2.4 |
| Distance from hospital (miles), median (IQR) | 14.9 (5.7, 37.1) | 16.1 (6.9, 49.2) | 22.2 |
| Income<sup>3</sup>, n (%) |  |  |  |
| <\$40,227 | 17 (7.8%) | 21 (9.7%) | 5.9 |
| \$40,227-\$50, 353 | 32 (14.7%) | 37 (17.1%) | 6.1 |
| \$50,354-\$62, 332 | 38 (17.5%) | 39 (18.0%) | 1.2 |
| ≥\$63,333 | 90 (41.5%) | 79 (36.4%) | -10.7 |
| Education<sup>4</sup>, n (%) |  |  |  |
| ≥17.6% | 19 (8.8%) | 23 (10.6%) | 5.6 |
| 10.9%-17.5% | 40 (18.4%) | 44 (20.3%) | 4.5 |
| 6.3%-10.8% | 57 (26.3%) | 54 (24.9%) | -3.2 |
| <6.3% | 61 (28.1%) | 55 (25.3%) | -6.6 |

<sup>1</sup>IQR, Interquartile Range
<sup>2</sup>CDCC, Charlson comorbidity score
<sup>3</sup>Income level is average household income of patient's home zip code
<sup>4</sup>Education is percentage of adults (age≥25 years) in the patient's zip code who did not graduate from high school

**Table 5:** Disease Characteristics for Patients who Received Neoadjuvant ChemoRT or ChemoRT+I/O – Propensity Matched Analysis.

|  | **ChemoRT**<br>**n=217** | **ChemoRT+I/O**<br>**n=217** | **Standardized**<br>**Differences** |
| --- | --- | --- | --- |
| Clinical T Status, n (%) |  |  |  |
| T1 | 3 (1.4%) | 5 (2.3%) | 4.9 |
| T2 | 54 (24.9%) | 46 (21.2%) | -8.9 |
| T3 | 157 (72.4%) | 159 (73.3%) | 2.1 |
| T4 | 3 (1.4%) | 7 (3.2%) | 11.8 |
| Clinical N Status, n (%) |  |  |  |
| N0 | 59 (27.2%) | 61 (28.1%) | 0.9 |
| N1 | 112 (51.6%) | 113 (52.1%) | 0.9 |
| N2 | 43 (19.8%) | 40 (18.4%) | -3.8 |
| N3 | 3 (1.4%) | 3 (1.4%) | 0.0 |
| Tumor location, n (%) |  |  |  |
| Upper third of esophagus | 1 (0.5%) | 1 (0.5%) | 0.0 |
| Middle third of esophagus | 13 (6.0%) | 13 (6.0%) | 0.0 |
| Lower third of esophagus | 185 (85.3%) | 188 (86.6%) | 3.7 |
| Overlapping lesion | 3 (1.4%) | 4 (1.8%) | 2.6 |
| Unspecified location | 15 (6.9%) | 11 (5.1%) | -9.0 |
| Histology, n (%) |  |  |  |
| Adenocarcinoma | 196 (90.3%) | 202 (93.1%) | 8.5 |
| Squamous cell | 16 (7.4%) | 12 (5.5%) | -6.2 |
| Adenosquamous | 2 (0.9%) | 1 (0.5%) | 0.0 |
| Bronchioloalveolar | 3 (1.4%) | 2 (0.9%) | -5.5 |
| Grade, n (%) |  |  |  |
| Well differentiated | 2 (0.9%) | 3 (1.4%) | 3.3 |
| Moderately differentiated | 71 (32.7%) | 69 (31.8%) | -2.1 |
| Poorly differentiated | 40 (18.4%) | 39 (18.0%) | -1.1 |
| Undifferentiated, anaplastic | 104 (47.9%) | 106 (48.8%) | 1.9 |
| Interval: chemotherapy to surgery (days), median (IQR) | 97.0 (84.0, 114.0) | 104.5 (87.0, 127.0) | 17.9 |
| Interval: radiation to surgery (days), median (IQR) | 95.0 (84.0, 111.0) | 95.5 (81.0, 111.0) | 0.2 |
| Operative Approach, n (%) |  |  |  |
| Open | 66 (30.4%) | 73 (33.6%) | 6.7 |
| VATS | 57 (26.3%) | 70 (32.3%) | 13.6 |
| Robot | 38 (17.5%) | 24 (11.1%) | -25.7 |

**Table 6:** Clinicopathologic Outcomes for Patients who Received Neoadjuvant ChemoRT or ChemoRT+I/O – Propensity Matched Analysis.

|  | **ChemoRT**<br>**(n=217)** | **ChemoRT+I/O**<br>**(n=217)** | **p-value** |
| --- | --- | --- | --- |
| Pathologic T Stage, n (%) |  |  | 0.72 |
| T0 | 25 (11.5%) | 25 (11.5%) | >.99 |
| T1 | 16 (7.4%) | 16 (7.4%) | >.99 |
| T2 | 24 (11.1%) | 16 (7.4%) | 0.14 |
| T3 | 35 (16.1%) | 35 (16.1%) | 0.90 |
| T4 | 0 (0.0%) | 1 (0.5%) | >.99 |
| Pathologic N Stage, n (%) |  |  | 0.84 |
| N0 | 69 (31.8%) | 57 (26.3%) | 0.17 |
| N1 | 28 (12.9%) | 26 (12.0%) | 0.88 |
| N2 | 9 (4.1%) | 10 (4.6%) | 0.82 |
| N3 | 2 (0.9%) | 3 (1.4%) | >.99 |
| Pathologic M Stage, n (%) |  |  | 0.45 |
| M0 | 0 (0.0%) | 1 (0.5%) | 0.32 |
| M1 | 217 (100.0%) | 216 (99.5%) | 0.85 |
| Complete Pathologic Response, n (%) | 21 (21%) | 20 (22%) | 0.90 |
| Nodal upstaging, n (%)<sup>1</sup> | 13 (13%) | 15 (16%) | 0.51 |
| Nodal downstaging from N2 to N0, n (%)<sup>2</sup> | 12 (27.9%) | 8 (20%) | 0.40 |
| Nodal downstaging from N2 to N1 or N0, n (%)<sup>2</sup> | 17 (39.5%) | 13 (32.5%) | 0.45 |
| Patients with lymph nodes examined, n (%) | 210 (96.8%) | 206 (94.9%) | 0.34 |
| Lymph nodes examined, median (IQR) | 17.0 (12.0, 24.0) | 17.0 (11.0, 26.0) | 0.98 |
| Surgical margins, n (%) |  |  | 0.65 |
| No residual tumor | 206 (94.9%) | 201 (92.6%) | 0.32 |
| Residual tumor, not specified | 5 (2.3%) | 5 (2.3%) | >.99 |
| Microscopic residual tumor | 4 (1.8%) | 4 (1.8%) | >.99 |
| Macroscopic residual tumor | 0 (0.0%) | 1 (0.5%) | >.99 |
| Unknown or not evaluable | 2 (0.9%) | 6 (2.8%) | 0.25 |
| Length of stay (days), median (IQR) | 9.0 (7.0, 13.0) | 9.0 (7.0, 13.0) | 0.60 |
| 30-day unplanned readmission | 6 (2.8%) | 8 (3.7%) | 0.40 |
| 30-day mortality, n (%) | 1 (0.5%) | 2 (0.9%) | 0.88 |
| 90-day mortality, n (%) | 11 (5.1%) | 8 (3.7%) | 0.73 |

<sup>1</sup>Denominator is the number with cN0 disease (chemoRT: n=59; chemoRT+I/O: n=61)
<sup>2</sup>Denominator is the number with cN2 disease (chemoRT: n=43; chemoRT+I/O: n = 40)

### COMMENT

In this study, we used national data to compare short- and intermediate-term clinical and oncologic outcomes among patients with locally advanced esophageal cancer who underwent esophagectomy following neoadjuvant chemoRT versus chemoRT+I/O. On unadjusted analysis, patients receiving neoadjuvant chemoRT appeared to have fewer lymph nodes examined and a greater rate of pathologic nodal downstaging. However, there were no differences in clinicopathologic outcomes including 30- and 90-day mortality between cohorts in unadjusted and propensity score-matched analyses. In unadjusted analysis, chemoRT+I/O patients had significantly higher 3-year OS compared to the chemoRT cohort, but these differences were not observed in propensity-score matched cohorts. These data expand a growing body of literature supporting the safety of neoadjuvant I/O to trimodality treatment for locally advanced esophageal cancer.

When transitioning promising approaches like chemoRT+I/O from the advanced to curative paradigm, evaluating safety is paramount. Recent phase III studies have described tolerability and efficacy of I/O when used in combination with chemotherapy in patients with unresectable or metastatic esophageal cancer^8,9^. Checkmate 577 demonstrated the disease-free survival benefit of adjuvant I/O after trimodality therapy^10^. In the neoadjuvant setting, there are limited data exploring safety and feasibility, and a greater paucity of data on oncologic outcomes. The phase II PERFECT trial of neoadjuvant CROSS-based chemoRT with atezolizumab versus chemoRT alone demonstrated feasibility without differences in pathologic complete response or overall survival rates^6^. In terms of surgical outcomes, Sihag et al. were the first to report post-esophagectomy outcomes comparing neoadjuvant chemoRT with durvalumab versus chemoRT alone showing comparable 30-day morbidity, readmissions, and mortality between cohorts^4^.

The results from our study are consistent with and extend from those of prior research. Paralleling Sihag et al., we found similar 30-day mortality rates among the neoadjuvant chemoRT+I/O patients, though lower 30-day readmission rates. The latter finding in our study is potentially associated with the younger age of the chemoRT+I/O cohort. We also observed that chemoRT+I/O patients appeared to have a greater number of lymph nodes examined, a potentially clinically meaningful finding given recent data suggesting a survival benefit following a more extensive lymphadenectomy^11,12,13^. This may be a result of I/O leading to pathologically enlarged lymph nodes, an observed phenomenon in I/O-treated non-small cell lung cancers^14^. Consistent with prior research, we found no differences in adjusted 30-day readmission and mortality rates between neoadjuvant chemoRT versus chemoRT+I/O patients. However, we also observed similar intermediate-term survival outcomes, such as 90-day mortality and 3-year overall survival between chemoRT and chemoRT+I/O cohorts. These data suggest that there may be sustained safety and comparable efficacy of chemoRT and chemoRT+I/O followed by esophagectomy.

Recently, Ge et al. reported the pooled results of a systemic review and meta-analysis of 27 clinical trials evaluating outcomes among patients treated with neoadjuvant systemic therapy inclusive of I/O followed by surgery^15^. The results demonstrated favorable pathologic complete and major response rates. However, the data did not include a neoadjuvant treatment modality inclusive of radiotherapy. Emerging data on the cellular effects of radiotherapy in the treatment of solid tumors has demonstrated upregulation of PD-L1 receptor expression on tumor cells as well as within the tumor microenvironment^16^. This suggests that there may be a synergistic mechanism in terms of anti-tumor effect following a neoadjuvant chemoRT+I/O in esophageal cancer. The findings presented in the current study do not include any data to examine the biologic mechanisms following each neoadjuvant regimen, nor adequate study sample to observe any differences between neoadjuvant chemotherapy+I/O versus chemoRT+I/O. Nevertheless, comparing various neoadjuvant treatment approaches to better understand the pathobiological mechanisms as well as oncologic outcomes represents a compelling area of future study.

We observed several factors associated with the addition of I/O to neoadjuvant chemoRT. Younger age was associated with the receipt of neoadjuvant chemoRT+I/O. In a recent NCDB analysis of patients receiving I/O for metastatic melanoma, Moyers et al. similarly identified younger age as predictor of receiving I/O, but also other important sociodemographic not uncovered in this analysis, including patient comorbidities, facility characteristics (e.g., academic centers) and payer/insurance type^17^. Further studies are needed to understand additional predictors of receipt of neoadjuvant chemoRT+I/O in esophageal cancer and, furthermore, whether these are associated with differential impact on outcomes. We also observed that a longer interval between chemotherapy and surgery was associated with the receipt of chemoRT+I/O. This observation may have biologic and oncologic impact, as existing data suggests a longer interval could allow for a more robust immune-mediated antitumor effect, maximizing potential for a pathologic response^18^. Lastly, patients with squamous cell histology were less likely to receive I/O as a component of neoadjuvant therapy. This observation likely reflects the differential response to chemoRT between adenocarcinoma and squamous cell carcinoma.

### Limitations

The results of this study must be interpreted in the context of several inherent limitations. This was a retrospective study harnessing data within the NCDB. While this allows for understanding the effectiveness of chemoRT versus chemoRT+I/O regimens, the results are subject to limitations of the database (e.g., external validity, confounding, or selection biases). To explore these sources of bias, we performed unadjusted, adjusted, and propensity score-matched analyses, each demonstrating similar findings. Our study was also constrained by the lack of data on surgical (e.g., type of esophagectomy performed, surgeon volume), clinic (e.g., specific postoperative complications) and I/O related data (e.g., specific I/O administered, tumor biology [PD-L1 expression, CPS score], or immune-related adverse events).

### Conclusion

This national analysis of neoadjuvant combined modality therapy (chemoRT+I/O versus chemoRT) for locally advanced esophageal cancer demonstrated no differences in short and intermediate clinical outcomes. These data support the growing body of literature demonstrating the safety and feasibility of adding I/O to chemoRT-based neoadjuvant regimens. As chemoRT+I/O combination approaches are expanded, further studies are needed to evaluate patient- and disease-related factors driving the patterns of clinical implementation, as well as long-term survival benefits.

## Data Availability

All data produced in the present study are available upon reasonable request to the authors
All data produced in the present work are contained in the manuscript
All data produced are available online at https://www.dropbox.com/scl/fo/2wyeihroquc88a09ftvgw/h?rlkey=brt8cumkb3yo87rs8d87bqcty&dl=0

## REFERENCES

1. van Hagen P, Hulshof MCCM, van Lanschot JJB, et al. Preoperative Chemoradiotherapy for Esophageal or Junctional Cancer. N Engl J Med. 2012;366(22):2074–2084. doi:10.1056/NEJMoa1112088

2. Blum Murphy M, Xiao L, Patel VR, et al. Pathological complete response in patients with esophageal cancer after the trimodality approach: The association with baseline variables and survival—The University of Texas MD Anderson Cancer Center experience. Cancer. 2017;123(21):4106–4113. doi:10.1002/cncr.30953

3. Klebanoff CA, Gattinoni L, Restifo NP. CD8+ T-cell memory in tumor immunology and immunotherapy. Immunol Rev. 2006;211:214–224. doi:10.1111/j.0105-2896.2006.00391.x

4. Sihag S, Ku GY, Tan KS, et al. Safety and feasibility of esophagectomy following combined immunotherapy and chemoradiotherapy for esophageal cancer. J Thorac Cardiovasc Surg. 2021;161(3):836–843.e1. doi:10.1016/j.jtcvs.2020.11.106

5. Yamamoto S, Kato K, Daiko H, et al. FRONTiER: A feasibility trial of nivolumab with neoadjuvant CF or DCF therapy for locally advanced esophageal carcinoma (JCOG1804E)—The short-term results of cohort A and B. JCO. 2021;39(3_suppl):202–202. doi:10.1200/JCO.2021.39.3_suppl.202

6. van den Ende T, de Clercq NC, van Berge Henegouwen MI, et al. Neoadjuvant Chemoradiotherapy Combined with Atezolizumab for Resectable Esophageal Adenocarcinoma: A Single-arm Phase II Feasibility Trial (PERFECT). Clin Cancer Res. 2021;27(12):3351–3359. doi:10.1158/1078-0432.CCR-20-4443

7. Ho D, Imai K, King G, Stuart EA. MatchIt: Nonparametric Preprocessing for Parametric Causal Inference. Journal of Statistical Software. 2011;42:1–28. doi:10.18637/jss.v042.i08

8. Sun JM, Shen L, Shah MA, et al. Pembrolizumab plus chemotherapy versus chemotherapy alone for first-line treatment of advanced oesophageal cancer (KEYNOTE-590): a randomised, placebo-controlled, phase 3 study. The Lancet. 2021;398(10302):759–771. doi:10.1016/S0140-6736(21)01234-4

9. Luo H, Lu J, Bai Y, et al. Effect of Camrelizumab vs Placebo Added to Chemotherapy on Survival and Progression-Free Survival in Patients With Advanced or Metastatic Esophageal Squamous Cell Carcinoma: The ESCORT-1st Randomized Clinical Trial. JAMA. 2021;326(10):916–925. doi:10.1001/jama.2021.12836

10. Kelly RJ, Ajani JA, Kuzdzal J, et al. Adjuvant Nivolumab in Resected Esophageal or Gastroesophageal Junction Cancer. New England Journal of Medicine. 2021;384(13):1191–1203.

11. Raja S, Rice TW, Murthy SC, et al. Value of Lymphadenectomy in Patients Receiving Neoadjuvant Therapy for Esophageal Adenocarcinoma. Ann Surg. 2021;274(4):e320–e327. doi:10.1097/SLA.0000000000003598

12. Samson P, Puri V, Broderick S, Patterson GA, Meyers B, Crabtree T. Extent of Lymphadenectomy Is Associated With Improved Overall Survival After Esophagectomy With or Without Induction Therapy. Ann Thorac Surg. 2017;103(2):406–415. doi:10.1016/j.athoracsur.2016.08.010

13. Panda N, Schumacher L. Surgical Approach to Esophagectomy Post CheckMate 577: Do Lymph Nodes Matter if Everyone Gets Adjuvant Immunotherapy? Thorac Surg Clin. 2023;33(2):209–213. doi:10.1016/j.thorsurg.2023.01.002

14. Cascone T, Weissferdt A, Godoy MCB, et al. Nodal immune flare mimics nodal disease progression following neoadjuvant immune checkpoint inhibitors in non-small cell lung cancer. Nat Commun. 2021;12(1):5045. doi:10.1038/s41467-021-25188-0

15. Ge F, Huo Z, Cai X, et al. Evaluation of Clinical and Safety Outcomes of Neoadjuvant Immunotherapy Combined With Chemotherapy for Patients With Resectable Esophageal Cancer: A Systematic Review and Meta-analysis. JAMA Network Open. 2022;5(11):e2239778. doi:10.1001/jamanetworkopen.2022.39778

16. Wang NH, Lei Z, Yang HN, et al. Radiation-induced PD-L1 expression in tumor and its microenvironment facilitates cancer-immune escape: a narrative review. Ann Transl Med. 2022;10(24):1406. doi:10.21037/atm-22-6049

17. Moyers JT, Patel A, Shih W, Nagaraj G. Association of Sociodemographic Factors With Immunotherapy Receipt for Metastatic Melanoma in the US. JAMA Network Open. 2020;3(9):e2015656. doi:10.1001/jamanetworkopen.2020.15656

18. Brahmer JR, Lacchetti C, Schneider BJ, et al. Management of Immune-Related Adverse Events in Patients Treated With Immune Checkpoint Inhibitor Therapy: American Society of Clinical Oncology Clinical Practice Guideline. J Clin Oncol. 2018;36(17):1714–1768. doi:10.1200/JCO.2017.77.6385

