## Supplemental Tables for "Neoadjuvant Immunotherapy and Chemoradiation Followed by Esophagectomy for Esophageal Cancer": Supplementary_Tables_and_Figures.docx

**Supplementary Table 1. Factors Associated with Receipt of Neoadjuvant ChemoRT+I/O**

|  | **Odds Ratio** | **95% CI** | | ***p-*value** |
| --- | --- | --- | --- | --- |
| Age (per year) | 0.97 | 0.94 | 0.99 | 0.02 |
| Sex (ref=male) | 0.55 | 0.29 | 1.03 | 0.06 |
| Insurance Status (ref=Uninsured) |  |  |  |  |
| Private | 0.55 | 0.20 | 1.57 | 0.27 |
| Medicaid | 0.31 | 0.09 | 1.14 | 0.08 |
| Medicare | 0.50 | 0.17 | 1.50 | 0.22 |
| Other Government | 0.13 | 0.01 | 1.23 | 0.08 |
| Facility Type (ref=Community cancer program) |  |  |  |  |
| Comprehensive community | 0.61 | 0.33 | 1.13 | 0.12 |
| Academic/research program | 1.06 | 0.67 | 1.67 | 0.82 |
| Integrated network cancer program | 1.00 | 0.99 | 1.00 | 0.39 |
| Education^1^ (ref=>17.6%) |  |  |  |  |
| 10.9%-17.5% | 0.83 | 0.46 | 1.49 | 0.54 |
| 6.3%-10.8% | 1.06 | 0.61 | 1.84 | 0.85 |
| <6.3% | 1.53 | 0.87 | 2.67 | 0.14 |
| Tumor Location (ref=Upper third) |  |  |  |  |
| Middle third of esophagus | 0.79 | 0.10 | 6.53 | 0.83 |
| Lower third of esophagus | 0.99 | 0.13 | 7.49 | >.99 |
| Overlapping lesion | 0.53 | 0.05 | 5.43 | 0.60 |
| Unspecified location | 0.63 | 0.06 | 6.40 | 0.70 |
| Histology (ref=Adenocarcinoma) |  |  |  |  |
| Squamous cell | 0.43 | 0.20 | 0.91 | 0.03 |
| Large cell | 1.00 | 0.99 | 1.00 | 0.93 |
| Adenosquamous | 1.01 | 0.14 | 7.57 | >.99 |
| Bronchioloalveolar | 1.40 | 0.34 | 5.82 | 0.64 |
| Grade (ref=Well differentiated) |  |  |  |  |
| Moderately differentiated | 2.24 | 0.54 | 9.36 | 0.27 |
| Poorly differentiated | 1.01 | 0.24 | 4.33 | >.99 |
| Undifferentiated, anaplastic | 2.27 | 0.55 | 9.41 | 0.26 |
| Interval: chemotherapy to surgery (days), median (IQR) | 1.02 | 1.01 | 1.02 | <.001 |
| Operative Approach (ref=Open) |  |  |  |  |
| VATS | 1.36 | 0.93 | 1.98 | 0.12 |
| Robot | 0.81 | 0.48 | 1.39 | 0.46 |

^1^Education is percentage of adults (age≥25 years) in the patient’s zip code who did not graduate from high school

**Supplementary Table 2. Cox Proportional Hazards Analysis Evaluating Overall Survival for Patients who Received Neoadjuvant ChemoRT or ChemoRT+I/O**

|  | **Hazard Ratio (HR)** | **95% CI** | | ***p-*value** |
| --- | --- | --- | --- | --- |
| Age (per year) | 1.02 | 1.01 | 1.02 | <.001 |
| Sex (ref=male) | 0.81 | 0.74 | 0.89 | <.001 |
| Race (ref=White) |  |  |  |  |
| Black | 1.16 | 0.97 | 1.38 | 0.10 |
| Other | 0.91 | 0.74 | 1.11 | 0.33 |
| CDCC score (ref=0)^1^ |  |  |  |  |
| 1 | 1.18 | 1.10 | 1.27 | <.001 |
| 2 | 1.30 | 1.15 | 1.47 | <.001 |
| 3+ | 1.34 | 1.13 | 1.59 | <.001 |
| Clinical T Stage (ref=T0) |  |  |  |  |
| T1 | 2.82 | 0.39 | 20.20 | 0.30 |
| T2 | 2.61 | 0.37 | 18.60 | 0.34 |
| T3 | 3.12 | 0.44 | 22.21 | 0.26 |
| T4 | 3.36 | 0.47 | 24.22 | 0.23 |
| Clinical N Stage (ref=N0) |  |  |  |  |
| N1 | 1.22 | 1.14 | 1.31 | <.001 |
| N2 | 1.24 | 1.13 | 1.37 | <.001 |
| N3 | 1.53 | 1.26 | 1.86 | <.001 |
| Insurance Status (ref=Uninsured) |  |  |  |  |
| Private | 0.72 | 0.57 | 0.90 | 0.01 |
| Medicaid | 0.90 | 0.70 | 1.16 | 0.40 |
| Medicare | 0.73 | 0.58 | 0.92 | 0.01 |
| Other Government | 0.83 | 0.61 | 1.12 | 0.22 |
| Insurance Status Unknown | 0.72 | 0.57 | 0.90 | 0.01 |
| Facility Type (ref=Community cancer program) |  |  |  |  |
| Comprehensive community | 0.81 | 0.63 | 1.05 | 0.12 |
| Academic/research program | 0.71 | 0.55 | 0.92 | 0.01 |
| Integrated network cancer program | 0.78 | 0.60 | 1.01 | 0.06 |
| Income^2^ (ref=<$46,227) |  |  |  |  |
| $46,227-$57,856 | 0.96 | 0.86 | 1.06 | 0.40 |
| $57,857-$74,062 | 0.87 | 0.78 | 0.97 | 0.01 |
| ≥$74,063 | 0.87 | 0.78 | 0.98 | 0.02 |
| Education^3^ (ref =>15.3%) |  |  |  |  |
| 9.1%-15.2% | 1.02 | 0.92 | 1.12 | 0.75 |
| 5.0%-9.0% | 1.03 | 0.93 | 1.15 | 0.53 |
| <5.0% | 0.98 | 0.87 | 1.11 | 0.78 |
| Grade (ref=Well differentiated) |  |  |  |  |
| Moderately differentiated | 1.17 | 0.98 | 1.39 | 0.08 |
| Poorly differentiated | 1.56 | 1.31 | 1.85 | <.001 |
| Undifferentiated, anaplastic | 1.32 | 1.09 | 1.60 | 0.01 |
| Tumor Location (ref=Upper third) |  |  |  |  |
| Middle third of esophagus | 1.15 | 0.82 | 1.60 | 0.42 |
| Lower third of esophagus | 0.94 | 0.68 | 1.31 | 0.72 |
| Overlapping lesion | 1.14 | 0.80 | 1.63 | 0.47 |
| Unspecified location | 0.98 | 0.68 | 1.41 | 0.92 |
| Histology (ref=Adenocarcinoma) |  |  |  |  |
| Squamous cell | 0.89 | 0.80 | 0.99 | 0.04 |
| Large cell | 1.03 | 0.64 | 1.63 | 0.92 |
| Adenosquamous | 1.13 | 0.80 | 1.61 | 0.48 |
| Bronchioloalveolar | 1.23 | 0.97 | 1.56 | 0.09 |
| Neoadjuvant ChemoRT+I/O | 1.04 | 0.65 | 1.65 | 0.88 |
| Year of diagnosis | 0.97 | 0.95 | 0.99 | <.001 |
| Distance from hospital (miles) | 1.00 | 1.00 | 1.00 | 0.58 |
| Operative Approach (ref=Open) |  |  |  |  |
| VATS | 0.95 | 0.88 | 1.01 | 0.11 |
| Robot | 0.88 | 0.80 | 0.97 | 0.01 |
